# Effect of Alert Level 4 on R_eff_: review of international COVID-19 cases

**DOI:** 10.1101/2020.04.30.20086934

**Authors:** Rachelle N. Binny, Shaun C. Hendy, Alex James, Audrey Lustig, Michael J. Plank, Nicholas Steyn

## Abstract

The effective reproduction number, *R_eff_*, is an important measure of transmission potential in the modelling of epidemics. It measures the average number of people that will be infected by a single contagious individual. A value of *R_eff_* > 1 suggests that an outbreak will occur, while *R_eff_* < 1 suggests the virus will die out. In response to the COVID-19 pandemic, countries worldwide are implementing a range of intervention measures, such as population-wide social distancing and case isolation, with the goal of reducing *R_eff_* to values below one, to slow or eliminate transmission. We analyse case data from 25 international locations to estimate their *R_eff_* values over time and to assess the effectiveness of interventions, equivalent to New Zealand’s Alert Levels 1-4, for reducing transmission. Our results show that strong interventions, equivalent to NZ’s Alert Level 3 or 4, have been successful at reducing *R_eff_* below the threshold for outbreak. In general, countries that implemented strong interventions earlier in their outbreak have managed to maintain case numbers at lower levels. These estimates provide indicative ranges of *R_eff_* for each Alert Level, to inform parameters in models of COVID-19 spread under different intervention scenarios in New Zealand and worldwide. Predictions from such models are important for informing policy and decisions on intervention timing and stringency during the pandemic.

**Executive Summary:**

- In response to the COVID-19 pandemic, countries around the world are implementing a range of intervention measures, such as population-wide social distancing and case isolation, with the goal of reducing the spread of the virus.
- *R_eff_*, the effective reproduction number, measures the average number of people that will be infected by a single contagious individual. A value of *R_eff_* > 1 suggests that an outbreak will occur, while *R_eff_* < 1 suggests the virus will die out.
- Comparing *R_eff_* in an early outbreak phase (no or low-level interventions implemented) with a later phase (moderate to high interventions) indicates how effective these measures are for reducing *R_eff_*.
- We estimate early-phase and late-phase *R_eff_* values for COVID-19 outbreaks in 25 countries (or provinces/states). Results suggest interventions equivalent to NZ’s Alert Level 3-4 can successfully reduce *R_eff_* below the threshold for outbreak.

## Introduction

On 11 March 2020, the World Health Organisation declared the outbreak of coronavirus (SARS-CoV-2) a pandemic. COVID-19, the disease caused by the coronovirus, was first detected in Wuhan, China in December 2019, and recent estimates (as of 3 March 2020) place its case fatality rate at 3.4% (ranging from 0.2 - 14.8%, depending on age and comorbidities) (WHO, 2020).

A key parameter in epidemiological modelling is the basic reproduction number, *R_0_*, which measures the average number of people that will be infected by a single contagious individual in a fully susceptible population. A value of *R_0_* > 1 indicates that an outbreak will occur, while *R_0_* < 1 means the virus will die out. During the early phase of an outbreak, the number of infected individuals in a population increases exponentially. As the virus spreads and more people become infected, the number of susceptible individuals (those not yet exposed to the virus) declines and this slows the rate of increase due to herd immunity. Eventually the number of infected individuals reaches a peak and then declines. Larger values of *R_0_* > 1 are associated with a faster increase in the number of infected individuals in a population during the early phase of an outbreak, a higher peak in daily case numbers, and a larger number of cumulative cases at the end of the outbreak.

Interventions, such as population-wide social distancing and case isolation, slow the spread of the virus by reducing the number of contacts between infectious and susceptible individuals. This leads to a reduction in the effective reproduction number, which we denote *R_eff_* to distinguish it from the basic reproduction number *R_0_* in the absence of control measures. The reduction in *R_eff_* slows the rate of increase in the early phase and results in a lower peak in the number of cases.

If a suppression strategy is employed, the aim of interventions is to reduce *R_eff_* to values less than one, to reduce the number of cases to very low levels. A mitigation strategy also aims to reduce *R_eff_* but to values *R_eff_ >1*, thereby ‘flattening the curve’ but not entirely preventing transmission. Recent studies have modelled these strategies to assess their feasability and implications for healthcare systems and fatality rates in the context of COVID-19 (Ferguson *et al*. 2020; James *et al*. 2020). Therefore, *R_eff_* can be a useful measure to inform policy and decision making on when interventions should be implemented or lifted, and what strength of intervention is required to achieve a desired reduction in transmission.

*R_eff_* has been reported for many countries experiencing COVID-19 outbreaks worlwide (see e.g. Abbott et al, 2020). Prior to interventions being implemented and assuming a population entirely susceptible to infection, *R_0_* values ranging from approximately 1.5 to 6 have been reported for COVID-19 (Abbott et al, 2020; Alimohamadi et al, 2020; Flaxman et al, 2020). These values indicate outbreaks would likely spread to a large proportion of the population in the absence of control measures. *R_eff_* estimates for later phases of outbreaks, after interventions aimed at mitigation or suppression were implemented, are reduced to values ranging from approximately 0.4 to 2 (Abbott et al, 2020; Flaxman et al, 2020; Price et al, 2020). However, the observed reduction in *R_eff_* varies widely between countries due to differences in the speed and stringency with which intervention measures were implemented, as well as other social and political factors.

To date, obtaining reliable *R_eff_* estimates for Aotearoa New Zealand has been challenging as case numbers have remained relatively low and dominated by imported international cases. In addition, we have not spent sufficient time under Alert Levels 1-3 to reliably assess their effects. However, assessing changes in *R_eff_* observed internationally and aligning these with the timings/strength of interventions viewed in the context of New Zealand’s Alert Level system, gives an indication of what could be expected in New Zealand. We review trajectories of numbers of new confirmed COVID-19 cases and deaths from 28 January 2020 to 17 April 2020, reported by 25 locations (countries or states/provinces), to estimate their *R_eff_* values. We compare *R_eff_* estimates in an early outbreak phase (typically no or low-level interventions implemented) with a later phase (moderate or high control interventions in place) to assess how effective interventions, equivalent to New Zealand’s Alert Levels 1-4, have been at reducing transmissibility of the virus. These values will be useful for informing future modelling in the New Zealand context (see e.g. Plank et al, 2020) and estimates will be updated as new data become available.

## Methods

We analysed data on numbers of confirmed cases and deaths over time (the ‘epidemic curve’) from 25 locations (states/provinces or countries) experiencing an outbreak of COVID-19 between 28/01/2020 and 17/04/2020, to assess the effective reproduction number (*R_eff_*) and efficacy of interventions at reducing *R_eff_*. Data were sourced from the data repository for the 2019 Novel Coronavirus Visual Dashboard, provided by the Johns Hopkins University (JHU) Center for Systems Science and Engineering (available from: https://github.com/CSSEGISandData/COVID-19). Data in the repository were compiled by JHU from multiple sources including the World Health Organisation (full source list in the provided link) and are updated daily. Confirmed cases include presumptive positive cases and, for the USA, death totals include those classified as confirmed and probable. Locations with highest cumulative case totals were selected for analysis provided there was sufficient information available on implemented interventions. We also included locations that were of particular interest due to, for example, their testing/reporting protocols or intervention approach, so long as they had reported at least 1000 cumulative cases. Data on intervention measures was obtained for each location using the International Monetary Fund’s Policy Tracker (https://www.imf.org/en/Topics/imf-and-covid19/Policy-Responses-to-COVID-19#N), supplemented by information from other sources. We broadly grouped intervention measures into four levels of increasing intensity, equivalent to New Zealand’s Alert Level framework (available from: https://covid19.govt.nz/alert-system/covid-19-alert-system/#covid-19-alert-system): Level 1 – Prepare, Level 2 – Reduce, Level 3 – Restrict, and Level 4 – Eliminate. For each location we recorded the date on which each Alert Level was implemented.

When a virus initially emerges in a country, prior to an outbreak becoming fully established, daily new case numbers are relatively low and dominated by imported international cases. The epidemic curve typically grows erratically and at a sub-exponential rate (Chowell et al, 2016). After 100 cases the outbreak is likely to be established and driven mainly by community transmission (Mishra & Mishra, 2020). Therefore to obtain reliable estimates with our analytical approach we assume an outbreak is established from the 100th confirmed case of infection and report time in days since the 100th case. During the early phase of an established outbreak, the epidemic curve grows approximately exponentially in proportion to *e^rt^*. The exponential growth rate *r* provides an important measure of the outbreak’s severity and can be used to obtain an estimate of *R_eff_*. We therefore first fitted an exponential growth curve to i) the number of new confirmed cases per day (i.e. the incidence curve), and ii) the number of deaths per day, to estimate *r* and then calculated an associated *R_eff_* for each location (Ma, 2020).

We estimated a time-varying *r* using the number of new confirmed cases (or deaths) *c_i_* on each day *t_i_* in a 10-day sliding window, starting from the date of the 100th confirmed case (day 0 to day 9, inclusive). A linear least-squares estimate of *r*was obtained by fitting the linear regression ln (*c_i_*) = ln(*c_0_*) + *r t_i_*. This 10-day window was shifted forward sequentially in one-day increments and *r* re-estimated for each new window in the sequence. Trends in *r* estimates as the window moves from the early-phase exponential growth (typically no or low-level interventions) into a later phase (variable levels of interventions across different countries), give an indication of how effective interventions have been in reducing transmissibility of COVID-19. Doubling time *T_d_* is related to *r* by *T_d_* = ln(2)/*r*.

From each time-varying growth rate *r*, we obtained an estimate of the effective reproduction number *R_eff_* using the non-parametric approach of Wallinga & Lipsitch (2006), which uses the distribution of generation time (the time between an individual becoming infected and infecting another individual) to infer *R_eff_* rather than assume any particular S(E)IR model form. We used a generation time distribution, *w(a)*, where *a* is the infection age (i.e. time since infection), inferred directly from COVID-19 source-recipient data in March 2020 by Ferreti *et al*. (2020); a Weibull distribution with mean and median 5.0 days and standard deviation of 1.9 days (Weibull shape = 2.826; scale = 5.665). Wallinga & Lipsitch (2006) relate *R_eff_* to rby *R_eff_* = 1/*M*(-*r*), where 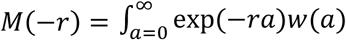 is the moment generating function of the generation time distribution, *w*(*a*). This relationship is derived using the Lotka-Euler equation, a popular theory in age-structured population growth modelling, under the assumption of a randomly mixed population undergoing exponential population growth and an age distribution that is stationary over time.

For ease of reporting, we selected a single 10-day window from the early phase (typically 0 – 9 days) and another from the late phase (i.e. after implementation of interventions) (day ranges of window varied among countries; see Appendix Table A1) to compare their *R_eff_* values (Fig. 1). However, figures showing trends in time-varying *R_eff_* over all time are provided in the Appendix. Confidence intervals are calculated for *R_eff_* under the assumption that the parameters of the generation time distribution are known, fixed and not subject to uncertainty.

**Figure 1.**
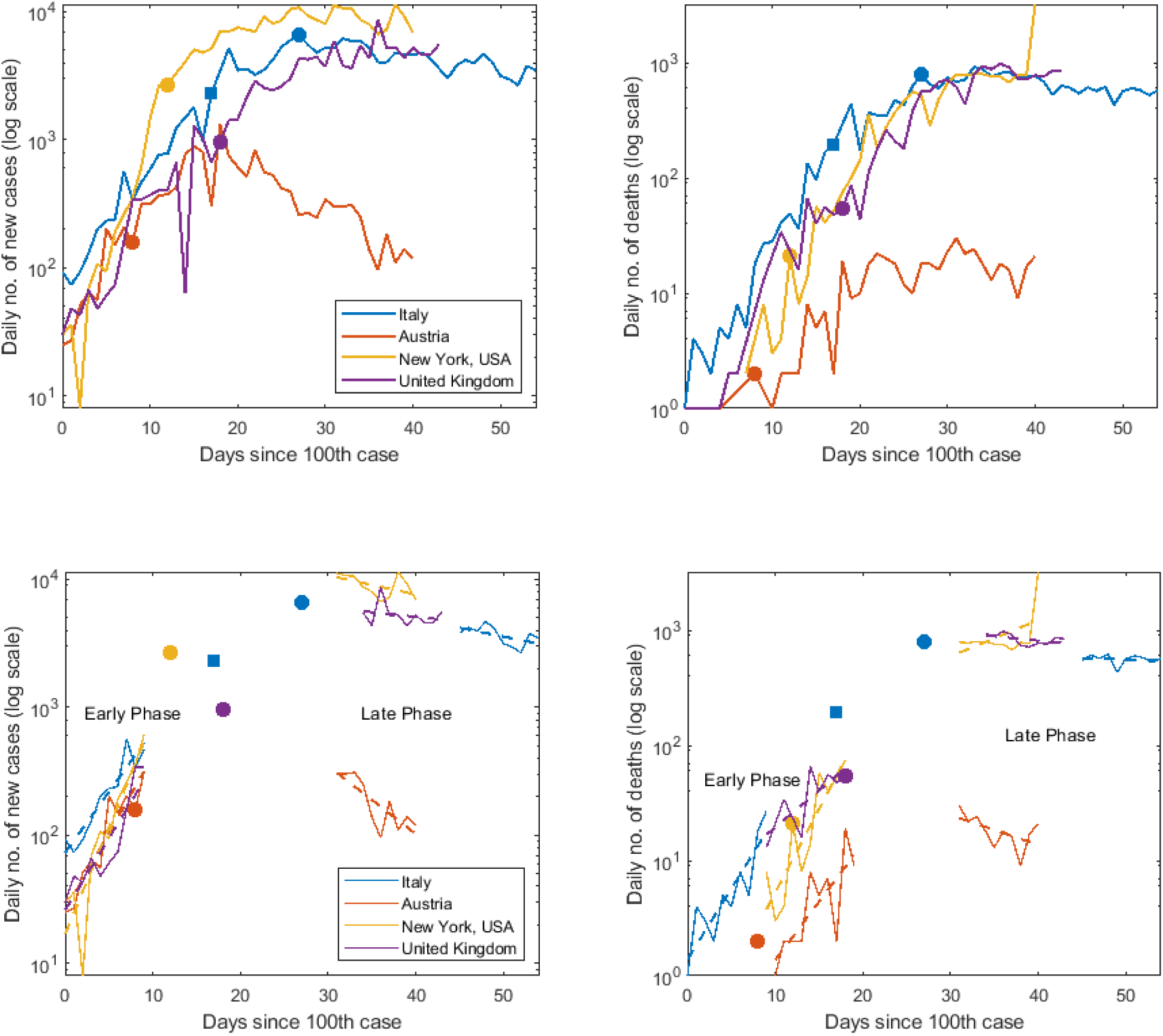
COVID-19 cases grow exponentially, however growth rates differ between countries, and between early and late phases. **Top row**: Daily numbers of new confirmed COVID-19 cases (left column) and new deaths (right column) since 100th confirmed case for outbreaks in four countries. Days on which Alert Level 3 (squared) and Alert Level 4 (circles) control measures were implemented. **Bottom row**: Exponential growth curves (dashed lines) fitted to 10 day windows of data (solid lines) in both the early phase and late phase.

Our approach has a number of limitations:

- Countries differ in their case definitions and testing/reporting protocols, making direct comparison of estimates difficult. We do not account for reporting delays or imported cases.
- Interventions implemented in other countries do not all align perfectly with New Zealand’s four Alert Levels. We provide details of key differences, however other differences (e.g. the speed and stringency at which interventions were implemented) will likely introduce additional variation between countries.
- Our estimates of *R_eff_* are dependent on the shape of the chosen generation time distribution. For simplicity, we assume parameters of this generation time distribution are known, fixed over time, and not subject to uncertainty.
- Asymptomatic individuals are not included in reported cases, however they still have the potential to contribute to transmission. Therefore, transmissibility could be under-estimated with this approach.
- Heterogeneity in transmission (e.g. effect of super-spreaders in the population) is not well captured by *R_eff_*.

Nonetheless, with these caveats in mind, our approach gives an indication of the reductions in *R_eff_* that have been achieved by interventions in other countries and frames these values in the context of New Zealand’s Alert Level system.

## Results

Comparison of *R_eff_* estimates in early and late phases suggests that interventions have reduced *R_eff_*, and that strong interventions (Alert Levels 3-4) can reduce *R_eff_* to values below the threshold for outbreak (*R_eff_* < 1) (Table 1, Fig. 1–2). However, the effectiveness of interventions for reducing *R_eff_* varies considerably between countries. Fitting to daily number of confirmed cases for 25 locations, yielded early-phase *R_eff_* estimates which were all greater than 1 (indicating outbreak), ranging from 1.5 to 5.4 (Table 1, Fig. 2). This range corresponds to doubling times of 8.7 days to 1.8 days. *R_eff_* values decreased in the late phase for 24 out of 25 locations, the exception being Singapore, where *R_eff_* declined over the first 30 days before increasing back up to early-phase transmission levels. Late-phase *R_eff_* estimates ranged from 0.3 to 2.1, and sixteen countries have reduced *R_eff_* to values less than 1 by implementing Alert Level 2-4 interventions. Twelve locations had sufficient data on daily number of deaths to allow estimation (Table 2). Early-phase *R_eff_* estimates ranged from 2.0 to 4.7 and decreased in the late phase for all twelve locations, with five locations achieving *R_eff_<* 1 through Alert Levels 3-4.

**Figure 2.**
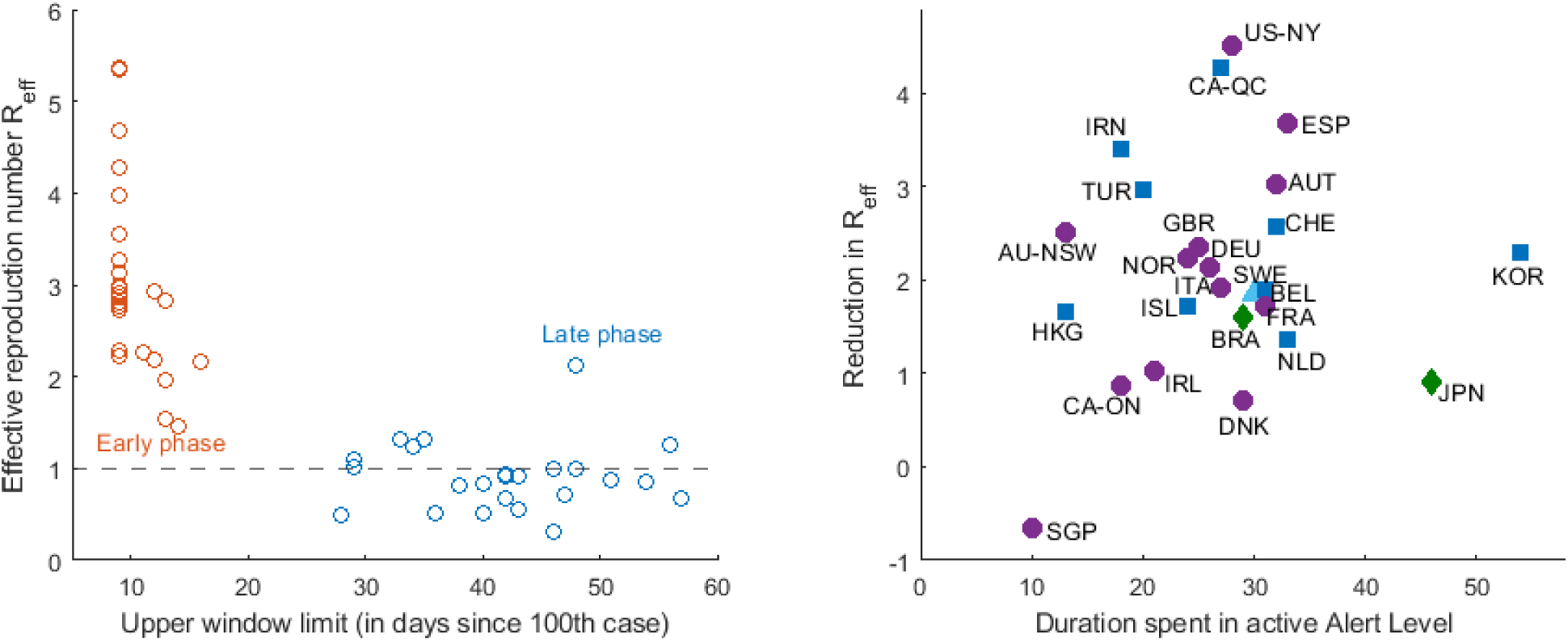
Interventions have successfully reduced *R_eff_* in most countries. Greatest reductions are observed in locations with longer durations (> 25 days) of sustained Alert Level 3-4 interventions. **Left**: *R_eff_* estimates for early phase of outbreak (red) (fitted to case data in a 10 day window, typically 0 - 9 days) and late phase (blue) (fitted to data in a 10-day window at the end of the time-series). Threshold for outbreak *R_eff_*=1 indicated by black dashed line. **Right**: Reduction in *R_eff_* (difference of early-phase and late-phase *R_eff_*) over varying durations of time spent in active Alert Level (days), for Level 1 (green diamonds), Level 2 (light blue triangles), Level 3 (dark blue squares) and Level 4 (purple circles) interventions.

**Table 1.** *R_eff_* estimates and associated exponential growth rates *r* (90% confidence intervals in parentheses), fitted to daily number of new cases. Estimates given for early and late phases (see Appendix Table A1) of COVID-19 outbreak in 25 countries/provinces/states, alongside start dates and durations that active control interventions have been in place up to 17 April 2020. Interventions were categorised into Alert Levels 1 to 4 of the New Zealand framework (L1 = Level 1, Prepare; L2 = Level 2, Reduce; L3 = Level 3, Restrict; L4 = Level 4, Eliminate). Locations are grouped by active Alert Level and in order of increasing late-phase *R_eff_*. *R_eff_* values in bold text indicate countries with average *R_eff_* < 1 in late phase, however not all of these were statistically significant at the 10% level (as indicated by 90% confidence intervals overlapping 1).

| Location | Early-phase<br>*R*<sub>eff</sub> | Late-phase<br>*R*<sub>eff</sub> | %<br>reduc-<br>tion<br>*R*<sub>eff</sub> | Early-phase *r* | Late-phase *r* | Date of<br>100th case | Cum.<br>cases | Cum.<br>deaths | Day control started<br>(in days since 100<sup>th</sup> case) |  |  |  | Duration<br>in active<br>Alert<br>Level<br>(days) |
| --- | --- | --- | --- | --- | --- | --- | --- | --- | --- | --- | --- | --- | --- |
|  |  |  |  |  |  |  |  |  | L1 | L2 | L3 | L4 |  |
| Alert Level 4 |  |  |  |  |  |  |  |  |  |  |  |  |  |
| New Zealand* | - | - | - | - | - | 19/03/2020 | 746 | 11 | Prior | 2 | 4 | 6 | 23 |
| New South Wales,<br>Australia<sup>1</sup> | 3.00<br>(2.02,4.33) | **0.49**<br>**(0.41,0.59)** | 84 | 0.24<br>(0.15,0.33) | -0.13<br>(-0.16,-0.1) | 14/03/2020 | 2926 | 26 | 2 | 7 | 8 | 15 | 13 |
| Austria | 3.55<br>(2.69,4.61) | **0.52**<br>**(0.38,0.72)** | 85 | 0.28<br>(0.21,0.34) | -0.12<br>(-0.18,-0.06) | 8/03/2020 | 14,595 | 431 | 2 | 2 | 8 | 8 | 32 |
| Norway | 2.91<br>(1.57,5.06) | **0.67**<br>**(0.27,1.52)** | 77 | 0.23<br>(0.09,0.37) | -0.08<br>(-0.24,0.09) | 6/03/2020 | 6937 | 161 | 6 | 6 | 10 | 18 | 24 |
| Germany | 2.86<br>(1.84,4.3) | **0.72**<br>**(0.46,1.08)** | 75 | 0.23<br>(0.13,0.33) | -0.06<br>(-0.14,0.02) | 1/03/2020 | 141397 | 4352 | 10 | 12 | 13 | 21 | 26 |
| Denmark<sup>2</sup> | 1.54<br>(1.09,2.13) | **0.82**<br>**(0.66,1.02)** | 47 | 0.09<br>(0.02,0.16) | -0.04<br>(-0.08,0) | 9/03/2020 | 7073 | 336 | 3 | 4 | 4 | 9 | 29 |
| New York, USA | 5.35<br>(3.03,8.89) | **0.84**<br>**(0.69,1.02)** | 84 | 0.38<br>(0.24,0.53) | -0.03<br>(-0.07,0) | 8/03/2020 | 230,597 | 14,832 | 4 | 12 | 12 | 12 | 28 |
| Italy | 2.77<br>(2.22,3.44) | **0.85**<br>**(0.72,1)** | 69 | 0.22<br>(0.17,0.27) | -0.03<br>(-0.06,0) | 23/02/2020 | 172,434 | 22,745 | 7 | 10 | 17 | 27 | 27 |
| United Kingdom<sup>3</sup> | 3.28<br>(2.26,4.65) | **0.92**<br>**(0.74,1.14)** | 72 | 0.26<br>(0.17,0.35) | -0.02<br>(-0.06,0.03) | 5/03/2020 | 108,692 | 14,576 | 10 | 11 | 18 | 18 | 25 |
| France | 2.73<br>(1.32,5.16) | 1.00<br>(0.41,2.18) | 63 | 0.22<br>(0.06,0.37) | 0.00<br>(-0.16,0.16) | 29/02/2020 | 147,969 | 18,681 | 13 | 13 | 14 | 17 | 31 |
| Spain<sup>4</sup> | 4.68<br>(3.44,6.26) | 1.00<br>(0.69,1.41) | 79 | 0.35<br>(0.27,0.43) | 0.00<br>(-0.07,0.07) | 2/03/2020 | 190,839 | 20,002 | 8 | 13 | 13 | 13 | 33 |
| Ireland | 2.26<br>(1.52,3.28) | 1.24<br>(0.82,1.82) | 45 | 0.17<br>(0.09,0.26) | 0.04<br>(-0.04,0.12) | 14/03/2020 | 13,980 | 530 | Prior | 13 | 13 | 13 | 21 |
| Ontario, Canada<sup>5</sup> | 2.19<br>(1.85,2.59) | 1.31<br>(0.93,1.83) | 40 | 0.17<br>(0.13,0.2) | 0.06<br>(-0.01,0.13) | 15/03/2020 | 10,456 | 524 | Prior | 2 | 8 | 15 | 18 |
| Singapore<sup>6</sup> | 1.46<br>(1.04,2.01) | 2.12<br>(1.69,2.64) | -45 | 0.08<br>(0.01,0.15) | 0.16<br>(0.11,0.21) | 29/02/2020 | 5050 | 11 | 3 | 11 | 26 | 38 | 10 |
| **Alert Level 3** |  |  |  |  |  |  |  |  |  |  |  |  |  |
| Hong Kong, China<sup>6,7</sup> | 1.97<br>(1.08,3.4) | **0.31**<br>**(0.17,0.52)** | 84 | 0.14<br>(0.02,0.27) | -0.22<br>(-0.31,-0.12) | 2/03/2020 | 1021 | 4 | Prior | 23 | 33 | - | 13 |
| Iceland<sup>6,7</sup> | 2.23<br>(1.32,3.62) | **0.5**<br>**(0.33,0.76)** | 78 | 0.17<br>(0.06,0.28) | -0.13<br>(-0.21,-0.05) | 12/03/2020 | 1754 | 9 | Prior | 4 | 12 | - | 24 |
| Switzerland | 3.14<br>(1.83,5.12) | **0.56**<br>**(0.38,0.81)** | 82 | 0.25<br>(0.13,0.37) | -0.11<br>(-0.18,-0.04) | 5/03/2020 | 27,078 | 1327 | Prior | 7 | 11 | - | 32 |
| South Korea | 2.98<br>(2.25,3.88) | **0.68**<br>**(0.57,0.79)** | 77 | 0.24<br>(0.17,0.3) | -0.08<br>(-0.11,-0.05) | 20/02/2020 | 10,635 | 230 | Prior | Prior | 3 | - | 54 |
| Iran<sup>8</sup> | 4.27<br>(3.16,5.68) | **0.87**<br>**(0.81,0.93)** | 80 | 0.32<br>(0.25,0.4) | -0.03<br>(-0.04,-0.01) | 26/02/2020 | 79,494 | 4958 | Prior | 8 | 33 | - | 18 |
| Netherlands<sup>9</sup> | 2.28<br>(1.63,3.12) | **0.91**<br>**(0.73,1.11)** | 60 | 0.17<br>(0.1,0.25) | -0.02<br>(-0.06,0.02) | 6/03/2020 | 30,449 | 3459 | 6 | 6 | 9 | - | 33 |
| Belgium | 2.83<br>(1.82,4.26) | **0.93**<br>**(0.59,1.43)** | 67 | 0.22<br>(0.12,0.32) | -0.01<br>(-0.1,0.07) | 6/03/2020 | 36,138 | 5163 | 4 | 6 | 11 | - | 31 |
| Turkey<sup>10</sup> | 3.97<br>(2.87,5.4) | 1.01<br>(0.92,1.11) | 75 | 0.31<br>(0.23,0.39) | 0.00<br>(-0.02,0.02) | 18/03/2020 | 78,546 | 1769 | Prior | Prior | 9 | - | 20 |
| Quebec, Canada<sup>11</sup> | 5.37<br>(3.06,8.9) | 1.10<br>(0.9,1.34) | 80 | 0.38<br>(0.24,0.53) | 0.02<br>(-0.02,0.06) | 19/03/2020 | 16,798 | 688 | Prior | Prior | 2 | - | 27 |
| **Alert Level 2** |  |  |  |  |  |  |  |  |  |  |  |  |  |
| Sweden | 2.80<br>(1.41,5.15) | **0.94**<br>**(0.72,1.23)** | 66 | 0.22<br>(0.07,0.37) | -0.01<br>(-0.06,0.04) | 6/03/2020 | 13,216 | 1400 | 5 | 12 | - | - | 30 |
| **Alert Level 1** |  |  |  |  |  |  |  |  |  |  |  |  |  |
| Japan<sup>12</sup> | 2.16<br>(1.72,2.70) | 1.25<br>(0.8,1.91) | 42 | 0.16<br>(0.11,0.21) | 0.05<br>(-0.04,0.13) | 21/02/2020 | 9787 | 190 | 10 | - | - | - | 46 |
| Brazil<sup>13</sup> | 2.92<br>(1.81,4.53) | 1.32<br>(0.94,1.82) | 55 | 0.23<br>(0.12,0.34) | 0.06<br>(-0.01,0.12) | 13/03/2020 | 33,682 | 2141 | 6 | - | - | - | 29 |

\*Confirmed and probable domestic cases only. All times in days since 10th case.
<sup>1</sup>Active Alert Level classified as AL4, however interventions have been less stringent than NZ (no formal school closure policy, but many are closed).
<sup>2</sup>Under-reporting of cases suspected due to a change in criteria for testing on 11 March (2 days since 100<sup>th</sup> case).
<sup>3</sup>Late onset of testing so may have reached 100 cases earlier than reported.
<sup>4</sup>Majority of intervention response was not implemented at a national level until 15 March (13 days since 100<sup>th</sup> case).
<sup>5</sup>Steadily increased stringency of interventions, as recently as 3 April (19 days since 100<sup>th</sup> case).
<sup>6</sup>High testing rates, results less likely to be affected by under-reporting (e.g. Iceland implemented a mobile tracing app from 1 April).
<sup>7</sup>Low case numbers, case data exhibits relatively high levels of variation. Interpret results with caution.
<sup>8</sup>Allocation to Alert Level 3 is an approximation due to limited available information on control measures. Domestic travel ban implemented.
<sup>9</sup>Mild cases not tested from 12 March so under-reporting likely. Alert Level 3 stronger than NZ's in place from 23 March (17 days since 100<sup>th</sup> case).
<sup>10</sup>Alert Level 3 less stringent than NZ.
<sup>11</sup>Alert Level 3 more stringent than NZ.
<sup>12</sup>Precise dates and stringency of interventions unknown.
<sup>13</sup>Regional control efforts.

**Table 2:** *R*eff estimates and associated exponential growth rates r (90% confidence intervals in parentheses), fitted to daily number of new deaths. Estimates given for early and late phases (see Appendix Table A1) of COVID-19 outbreak in 12 countries/states, alongside start dates and durations that active control interventions have been in place up to 17 April 2020. Interventions were categorised into Alert Levels 1 to 4 of the New Zealand framework (L1 = Level 1, Prepare; L2 = Level 2, Reduce; L3 = Level 3, Restrict; L4 = Level 4, Eliminate). Locations are grouped by active Alert Level and in order of increasing late-phase *R_eff_*. *R_eff_* values in bold text indicate countries with average *R_eff_* < 1 in late phase, however not all of these were statistically significant at the 10% level (as indicated by 90% confidence intervals overlapping 1).

| Location | Early-phase<br>*R<sub>eff</sub>* | Late-phase<br>*R<sub>eff</sub>* | %<br>reduc-<br>tion<br>*R<sub>eff</sub>* | Early-phase *r* | Late-phase *r* | Date of<br>100th case | Cum.<br>cases | Cum.<br>deaths | Day control started<br>(in days since 100<sup>th</sup> case) |  |  |  | Duration<br>in active<br>Alert<br>Level<br>(days) |
| --- | --- | --- | --- | --- | --- | --- | --- | --- | --- | --- | --- | --- | --- |
|  |  |  |  |  |  |  |  |  | L1 | L2 | L3 | L4 |  |
| Alert Level 4 |  |  |  |  |  |  |  |  |  |  |  |  |  |
| Austria | 2.93<br>(1.52,5.28) | 0.73<br>(0.53,1) | 75 | 0.23<br>(0.09,0.38) | -0.06<br>(-0.12,0) | 8/03/2020 | 14,595 | 431 | 2 | 2 | 8 | 8 | 32 |
| Spain | 4.66<br>(1.68,10.89) | 0.91<br>(0.69,1.2) | 80 | 0.35<br>(0.11,0.59) | -0.02<br>(-0.07,0.04) | 2/03/2020 | 190,839 | 20,002 | 8 | 13 | 13 | 13 | 33 |
| United Kingdom | 2.02<br>(1.24,3.17) | 0.92<br>(0.82,1.02) | 54 | 0.15<br>(0.04,0.25) | -0.02<br>(-0.04,0) | 5/03/2020 | 108,692 | 14,576 | 10 | 11 | 18 | 18 | 25 |
| Italy | 3.61<br>(2.37,5.33) | 0.99<br>(0.88,1.11) | 73 | 0.28<br>(0.18,0.38) | 0.00<br>(-0.02,0.02) | 23/02/2020 | 172,434 | 22,745 | 7 | 10 | 17 | 27 | 27 |
| France | 3.49<br>(2.08,5.59) | 1.05<br>(0.71,1.51) | 70 | 0.27<br>(0.15,0.4) | 0.01<br>(-0.07,0.08) | 29/02/2020 | 147,969 | 18,681 | 13 | 13 | 14 | 17 | 31 |
| Germany | 2.89<br>(1.75,4.58) | 1.05<br>(0.6,1.76) | 64 | 0.23<br>(0.12,0.34) | 0.01<br>(-0.1,0.12) | 1/03/2020 | 141,397 | 4352 | 10 | 12 | 13 | 21 | 26 |
| New York, USA | 4.45<br>(2.72,6.96) | 1.43<br>(0.93,2.13) | 68 | 0.34<br>(0.22,0.46) | 0.07<br>(-0.01,0.16) | 8/03/2020 | 230,597 | 14,832 | 4 | 12 | 12 | 12 | 28 |
| Alert Level 3 |  |  |  |  |  |  |  |  |  |  |  |  |  |
| Iran | 2.05<br>(1.64,2.53) | 0.82<br>(0.77,0.87) | 60 | 0.15<br>(0.1,0.2) | -0.04<br>(-0.05,-0.03) | 26/02/2020 | 79,494 | 4958 | Prior | 8 | 33 | - | 18 |
| Belgium | 4.59<br>(2.74,7.32) | 1.09<br>(0.83,1.42) | 76 | 0.34<br>(0.22,0.47) | 0.02<br>(-0.04,0.07) | 6/03/2020 | 36,138 | 5163 | 4 | 6 | 11 | - | 31 |
| Netherlands | 3.84<br>(2.58,5.57) | 1.11<br>(0.84,1.44) | 71 | 0.30<br>(0.2,0.39) | 0.02<br>(-0.03,0.07) | 6/03/2020 | 30,449 | 3459 | 6 | 6 | 9 | - | 33 |
| **Turkey** | 3.33<br>(1.82,5.71) | 1.21<br>(1.15,1.27) | 64 | 0.26<br>(0.12,0.4) | 0.04<br>(0.03,0.05) | 18/03/2020 | 78,546 | 1769 | Prior | Prior | 9 | - | 20 |
| ***Alert Level 2*** |  |  |  |  |  |  |  |  |  |  |  |  |  |
| **Brazil** | 3.80<br>(3.04,4.71) | 1.50<br>(1.09,2.03) | 61 | 0.30<br>(0.24,0.35) | 0.08<br>(0.02,0.15) | 13/03/2020 | 33,682 | 2141 | 6 | - | - | - | 29 |

The time taken by countries to first implement interventions equivalent to NZ’s Level 1 (Prepare) ranged up to 13 days since the 100th case. More stringent interventions equivalent to Level 4 (Eliminate) were implemented after 8 to 38 days in 13 locations, and had not yet been implemented in the remaining 12 locations (Table 1). New Zealand implemented Alert Level 3 and 4 interventions earlier (at 4 days and 6 days since 10th confirmed domestic case, respectively) than all other countries considered (Table 1). Because countries implemented interventions at different times and, to date, have spent varying durations of time in their active Alert Level, it is difficult to directly compare late-phase *R_eff_* estimates between countries (e.g. interventions remain in place and, in some locations, may not have had sufficient time to achieve their potential full effect). However, Figure 2 gives an indication of how the reduction in *R_eff_* varies with duration spent at the active Alert Level. In particular, for countries in the relatively early stages of Alert Level 4, we expect late-phase *R_eff_* to further decline with each new day spent at that Level, and will continue to track and update these estimates as new daily case data become available. In general, largest reductions in late-phase *R_eff_* were achieved in locations (e.g. New York state, USA and Quebec, Canada) that had sustained Alert Level 3-4 control for durations of more than 25 days. Countries that reduced late-phase *R_eff_* to *R_eff_* <1, achieved this after a lag of at least 1 to 2.5 weeks (see figures in Appendix).

## Discussion

In response to the COVID-19 pandemic, a large number of countries worldwide have implemented or are planning to implement national-scale control interventions to slow or eliminate transmission of the virus. Our review of the extent to which interventions have successfully reduced *R_eff_*, a measure of transmissibility, informs future modelling to predict the outcomes we might expect from interventions employed in Aotearoa New Zealand. Our analysis of case data for COVID-19 in 25 countries/states found that *R_eff_* consistently decreased between the early and late phases of exponential growth in all locations considered.

Lasting immunity is yet to be confirmed for those who recover from COVID-19, therefore a mitigation strategy aimed at allowing controlled spread (i.e. small *R_eff_* values greater than 1) does not guarantee herd immunity will develop in the population. There is therefore value in achieving *R_eff_* < 1, and our results show that several countries implementing strong interventions have successfully reduced *R_eff_* below this outbreak threshold. The time taken to implement high intensity (Alert Levels 3-4) interventions and the stringency with which they were implemented varied considerably between countries, as did the resulting late-phase *R_eff_* estimates. However, in general, faster implementation and longer durations of sustained Alert Level 3-4 interventions yielded greatest reductions in *R_eff_*.

Testing approaches vary widely within and between countries, as well as over time, therefore reported cases exhibit high uncertainty. Daily reported deaths also vary due to differences in reporting protocols, healthcare systems and whether a country/region’s healthcare operating capacity was exceeded. This makes it challenging to directly compare *R_eff_* estimates between countries. For some locations, our *R_eff_* values estimated from daily case numbers differed from the estimates obtained using daily number of deaths. For instance, nine out of twelve countries had higher estimates of late-phase *R_eff_* fitted to daily deaths compared to daily cases (though these were not statistically significant). This is expected, due to the time lag between an individual being reported as infected and death, which means fitting to daily deaths over a particular window of time actually provides an estimate from an earlier time in the epidemic.

These differences may also indicate that cases are under-reported. For example, a recent study by Russell et al (2020) assessed the extent of under-reporting of symptomatic COVID-19 cases globally; six out of these nine countries had reported fewer than 10% of symptomatic cases. Our approach does not account for transmission by asymptomatic individuals, who can still spread the virus but are not reported in case numbers (Lavezzo et al, 2020). Additionally, the lag between an individual being exposed to the virus and being confirmed as infectious by testing, means the full extent of the reduction in *R_eff_* after interventions, will not be observable in case data until several days after interventions are begun. Our results indicate this lag could be at least 1-2.5 weeks (see results in Appendix).

Our results show reasonable agreement to other studies on *R_eff_* for COVID-19 (e.g.Abbott et al, 2020; Alimohamadi et al, 2020; Cowling et al, 2020; Flaxman et al, 2020; Price et al (2020)). For example, we obtained similar late-phase *R_eff_* estimates but higher early-phase estimates than a global study by Abbott et al. (2020), which used an approach based on Thompson et al (2019). Our approach differs from Abbott et al (2020) because we do not account for reporting delays, right censoring of confirmation dates, or imported cases. Our estimates therefore exhibit higher uncertainty and are likely inflated by imported cases, particularly in the early phase. These additional factors should be addressed before obtaining estimates from New Zealand case data. Our estimates of *R_eff_* from the exponential growth rate are based on an assumed generation time distribution, which has mean 5 days. This was fitted to estimates of the times between infection of 40 source-recipient case pairs (Ferreti et al, 2020) and is subject to uncertainty. Other groups have estimated longer mean generation times (Flaxman et al 2020; Li et al, 2020). This would result in our method returning higher estimates for *R_eff_* in the initial phase before interventions. However, it would have less of an effect on the relative reduction in *R_eff_* as a result of interventions. Two other recent international studies have examined the effects of control interventions on exponential growth rates (Hsiang *et al*., 2020) and *R_eff_* (Flaxman *et al*., 2020) for COVID-19. We have taken a simpler theoretical approach to calculate and compare growth rates and *R_eff_*, however our estimates show reasonable agreement to values estimated in both studies and agree with the general result that *R_eff_* is considerably reduced by strong (e.g. lockdown-level) interventions.

We expect late-phase *R_eff_* estimates to continue to decrease as interventions are sustained for longer time periods and will continue to update our estimates. At the time of writing, case numbers are too low to apply this approach to New Zealand data (though we continue to closely monitor daily domestic cases) and New Zealand has spent insufficient time in Alert Levels 1-3 to assess the effects. We acknowledge that political, societal, cultural and other differences between countries mean that the effects of interventions observed in this analysis may differ from effects in New Zealand. Additionally, interventions employed abroad did not always fit precisely within the New Zealand framework, contributing further variation in estimates for each Alert Level. Nonetheless, this work provides indicative ranges of *R_eff_* for each Alert Level, to inform parameters for models until reliable estimates are available for NZ. For example, our ranges of late-phase estimates inform parameters for effectiveness of control and *R_eff_* in the stochastic model of Plank et al (2020) of COVID-19 spread and effects of interventions in Aotearoa New Zealand (see e.g. Tables 2–3 in Plank et al, 2020). Predictions from such models are important for informing policy and decisions on intervention timing and stringency.

## Data Availability

This article does not present new data. Link to source of data used in analyses provided.

https://github.com/CSSEGISandData/COVID-19

## Acknowledgements

The authors thank Dr Matt Parry, Professor Nigel French, Dr Anya Mizdrak, Dr Fraser Morgan, Dr Markus Luczak-Roesch, and Dr Samik Datta for their useful comments on the manuscript.

## References

Abbott S, Hellewell J, Munday JD, et al. (19 April 2020). Temporal variation in transmission during the COVID-19 outbreak. Available from: https://epiforecasts.io/covid/ (accessed 20 April 2020).

Alimohamadi Y, Taghdir M and Sepandi M (20 Mar 2020). The Estimate of the Basic Reproduction Number for Novel Coronavirus disease (COVID-19): A Systematic Review and Meta-Analysis. Journal of Preventive Medicine and Public Health. Doi: https://doi.org/10.3961/jpmph.20.076

Chowell G, Sattenspiel L, Bansal S and Viboud C (2016). Mathematical models to characterize early epidemic growth: A review. Physics of life reviews, 18, 66–97. Doi: https://doi.org/10.10167j.plrev.2016.07.005

Cowling BJ, Ali ST, Ng TW, Tsang TK, Li JC, Fong MW, Liao Q Kwan MY, Lee SL, Chiu SS and Wu JT (17 Apr 2020). Impact assessment of non-pharmaceutical interventions against coronavirus disease 2019 and influenza in Hong Kong: an observational study. The Lancet Public Health. Doi: https://doi.org/10.1016/S2468-2667(20)30090-6

James A, Hendy SC, Plank MJ and Steyn N (2020). Suppression and mitigation strategies for control of COVID-19 in New Zealand. *MedRxiv* 2020.03.26.20044677, https://doi.org/10.1101/2020.03.26.20044677

Lavezzo E, Franchin E, Ciavarella C, et al (18 Apr 2020). Suppression of COVID-19 outbreak in the municipality of Vo, Italy. medRxiv 2020.04.17.20053157; doi: https://doi.org/10.1101/2020.04.17.20053157

Ma (2020). Estimating epidemic exponential growth rate and basic reproduction number. Infectious disease modelling, 5, 129–141.

Ferguson N, et al. (16 Mar 2020). Impact of non-pharmaceutical interventions (NPIs) to reduce COVID-19 mortality and healthcare demand (Report 9). Imperial College London, doi: https://doi.org/10.25561/77482

Ferretti L, Wymant C, Kendall M, et al. (31 Mar 2020). Quantifying SARS-CoV-2 transmission suggests epidemic control with digital contact tracing. Science, doi: https://doi.org/10.1126/science.abb6936

Flaxman S, Mishra S, Gandy A, et al. (30 Mar 2020). Estimating the number of infections and the impact of non-pharmaceutical interventions on COVID-19 in 11 European countries. Imperial College London, doi: https://doi.org/10.25561/77731

Hsiang S, Allen D, Annan-Phan S, et al. (31 Mar 2020). The Effect of Large-Scale Anti-Contagion Policies on the Coronavirus (COVID-19) Pandemic. medRxiv 2020.03.22.20040642; doi: https://doi.org/10.1101/2020.03.22.20040642

Li Q Guan X, Wu P, Wang X, Zhou L, Tong Y et al (2020). Early Transmission Dynamics in Wuhan, China, of Novel Coronavirus-Infected Pneumonia. New England Journal of Medicine, 382: 1199–1207.

Mishra & Mishra (26 March 2020). A deductive approach to modeling the spread of COVID-19. MedRxiv preprint, doi: https://doi.org/10.1101/2020.03.26.20044651

Plank MJ, Binny RN, Hendy SC, Lustig A, James A, Steyn N (9 April 2020). A stochastic model for COVID-19 spread and the effects of Alert Level 4 in Aotearoa New Zealand. MedRxiv preprint, doi: https://doi.org/10.1101/2020.04.08.20058743

Price DJ, Shearer FM, Meehan M, McBryde E, Golding N, McVernon J, McCaw JM (14 April 2020). Estimating the case detection rate and temporal variation in transmission of COVID-19 in Australia. Technical Report. Available from: https://www.doherty.edu.au/uploads/content_doc/EstimatingchangesinthetransmissionofCOVID-19April14-public-release.pdf (Accessed on 20 April 2020)

Russell TW, Hellewell J, Abbott S, et al. (14 April 2020). Using a delay-adjusted case fatality ratio to estimate under-reporting. Available from: https://cmmid.github.io/topics/covid19/severity/globalcfrestimates.html (accessed 20 April 2020).

Sanche S, Lin YT, Xu C, Romero-Severson E, Hengartner N and Ke R (July 2020). High contagiousness and rapid spread of severe acute respiratory syndrome coronavirus 2. Emerg. Infect. Dis, 26(7). Doi: https://doi.org/10.3201/eid2607.200282

Thompson R, Stockwin J, Gaalen Rv, et al. (2019). Improved inference of time-varying reproduction numbers during infectious disease outbreaks. Epidemics, 29, 100356. Doi: https://doi.org/10.1016/j.epidem.2019.100356

WHO (3 March 2020) Director-General’s opening remarks at the media briefing on COVID-19, World Health Organization. Last accessed from https://www.worldometers.info/coronavirus/coronavirus-death-rate/#ref-13 on 30 March 2020.

Wallinga J, & Lipsitch M (2006). How generation intervals shape the relationship between growth rates and reproductive numbers. Proceedings of the Royal Society B: Biological Sciences, 274, 599e604.

